# A Novel, Widespread Impurity in Mass-Compounded Tirzepatide/B12 Products: Patient Safety Implications

**DOI:** 10.64898/2026.03.09.26347818

**Authors:** Brad Jordan, Luke Arbogast, Matthew Clemens, Lihua Huang, Matthew Snyder

## Abstract

**Background:** Compounded versions of tirzepatide are widely available in the U.S. in the form of fixed-dose combinations of tirzepatide and various analogs of vitamin B12. These combinations are mass marketed in the U.S. and other countries as comparable to FDA-approved tirzepatide products even though they undergo no evaluation of their potency or impurity profiles.

**Research Design and Methods:** Samples of compounded tirzepatide combined with B12 obtained from various sources in the U.S. market were tested using various analytical methods. Samples were assessed for unacceptable levels of peptide-related impurities.

**Results:** Our testing identified a widespread and previously unidentified impurity in compounded tirzepatide-B12 products resulting from a chemical reaction between tirzepatide and certain analogs of B12.

**Conclusion:** Despite the presence of this impurity, these products continue to be mass marketed as “personalized” treatments. Our findings underscore the importance of testing and FDA approval before new drugs are marketed and highlights potential risks for patients associated with untested combinations. A novel impurity, present at substantial levels in compounded tirzepatide/B12 products, highlights risks inherent in marketing complex therapies outside the drug-approval framework. Although clinical effects of this impurity are unknown, the identification of a widespread impurity adds to the existing quality concerns presented by compounded tirzepatide.

## Introduction

The United States Food and Drug Administration’s (FDA) approval of tirzepatide marked a major advance in incretin-based therapy, offering dual glucose-dependent insulinotropic polypeptide (GIP) and glucagon-like peptide-1 (GLP-1) receptor agonism that improves glycemic control and supports weight reduction in patients with obesity or overweight with at least one weight-related comorbidity. Branded, FDA-approved tirzepatide products (Mounjaro® and Zepbound®) are manufactured under strict current Good Manufacturing Practices (cGMP) and undergo extensive pre-market testing to ensure safety, efficacy, and quality.

In the United States, compounding pharmacies, medical spas (medspas), telehealth networks, and others began manufacturing and selling large quantities of compounded tirzepatide products while the approved products were listed on FDA’s shortage list. Following FDA’s determinations in October and December 2024 that tirzepatide was no longer in shortage, many shifted to producing and selling compounded tirzepatide in fixed-dose combinations with vitamins and other additives, including analogs of vitamin B12 (e.g., methylcobalamin or hydroxocobalamin). The true scope of these practices is unknown because these products exist outside the federal regulatory framework for pharmaceuticals. Sellers of unapproved compounded tirzepatide products generally do not conduct pre-market testing, file product applications, register their establishments, disclose their products or volume of products, follow cGMP, track or trace their products, inform FDA when manufacturing problems occur, or report adverse events.

The mass compounding of prescription drugs outside the FDA drug-approval framework has long been associated with serious risks, including substandard drug potency, bacterial contamination, potentially harmful impurities, patient harm and even death [1]. Compounded drugs accounted for 45% of all drug product recalls in the United States between 2012 and 2021, despite accounting for less than 3% of prescriptions, and the predominant reason for recall among compounders was a lack of assurance of sterility [2]. As of December 2, 2025, the FDA’s Adverse Event Reporting System (FAERS) database reflected at least 455 reports of adverse events with use of compounded tirzepatide, including 339 serious adverse events and 8 deaths [3]. A recent analysis of the FAERS database concluded that compounded tirzepatide may be associated with significantly higher odds of adverse events, safety concerns and product quality issues as compared to FDA-approved tirzepatide [4]. For these and other reasons, FDA has cautioned that the use of compounded tirzepatide “can be risky for patients” [5].

We report here findings in compounded, fixed-dose combinations of tirzepatide and B12 that reveal significant deviations from the approved tirzepatide drug composition in the form of contamination with a previously uncharacterized impurity. This impurity was identified in ten total samples of compounded tirzepatide formulated with B12 obtained from multiple compounding pharmacies, medspas, and telehealth networks. The impurity results from a chemical interaction between tirzepatide and B12 and was present at levels up to 10% of total protein content in the samples tested. There are currently no data on the impact of this impurity on the safety or efficacy of tirzepatide, and its presence raises substantial concerns about the safety and effectiveness of these compounded products.

## Regulatory Background

For a fixed-dose combination product—one that contains two or more active ingredients—FDA generally requires that each component be evaluated individually and in combination to demonstrate that the overall effects are clinically appropriate and do not introduce unacceptable risks [6]. Sponsors must conduct preclinical pharmacology and toxicology studies, followed by clinical trials that compare the combination to its individual components and/or placebo. Manufacturing must adhere to cGMP, which requires exhaustive characterization of each component and the combination, including reporting of any impurity present at 0.05%, identification of any impurity present at 0.1%, and qualification of any impurity present at 0.15% [7]. Stability testing also must confirm that the combination remains safe and effective throughout its shelf life. Internationally, regulatory bodies such as the European Medicines Agency (EMA) and the International Council for Harmonisation (ICH) have similar expectations, emphasizing justification for the combination, interaction studies, and robust quality control.

Compounded combination drugs generally do not comply with these standards. Compounded combinations are not evaluated through clinical studies that assess the contributions of each component or the interactions between them. Pharmacy-compounded drugs also are not held to the same manufacturing or product purity standards and are not required to report, identity, or qualify the impurities present.

To assess the quality of compounded tirzepatide products combined with B12, we obtained samples of these products from various sources across the United States (U.S.), including compounding pharmacies, medspas, and telehealth networks. We report here the results of these analyses and highlight significant concerns for patient safety that are posed by unregulated, mass manufacturing of untested combination tirzepatide products.

## Methods

Samples of compounded tirzepatide combined with various analogs of vitamin B12 (e.g., hydroxocobalamin and methylcobalamin) were obtained from various sources across the U.S. Importantly, it has been reported that the methylcobalamin analog of B12 spontaneously converts to the hydroxocobalamin analog in aqueous solutions [8]. All samples were subjected to analysis by Ultra-High Performance Liquid Chromatography-Mass Spectrometry (UPLC-MS). New samples were subsequently prepared by mixing tirzepatide with hydroxocobalamin at a 1:1 molar ratio (consistent with the makeup of the samples tested by UPLC-MS) in phosphate buffered saline (PBS) at pH 7.4 to mimic physiological pH conditions. These samples were subjected to a suite of 1D and 2D Nuclear Magnetic Resonance (NMR) experiments designed to further probe the interaction between B12 and tirzepatide and to evaluate any possible impact on the structure of the tirzepatide molecule. Full experimental details as well as additional figures are available in the Supplementary.

## Results

Analysis of the samples indicated a wide range of potencies when compared to tirzepatide reference standard, with some samples demonstrating levels as low as 43% of the labeled value. Additionally, we identified the presence of a previously unknown impurity with a retention time of approximately 28 minutes (Figure 1) that was present in all the samples analyzed. MS analysis indicated that the impurity had a mass of approximately 6,138 Daltons, larger than tirzepatide alone (4,810.52 Daltons). The results suggested that the impurity consisted of a B12 molecule covalently or coordinately attached to tirzepatide, likely via a ligand substitution reaction to form a new tirzepatide–B12 adduct. The tirzepatide–B12 adduct remained intact under denaturing conditions and did not dissociate during electrospray ionization mass spectrometry, suggesting a stable association. The tirzepatide-B12 adduct was present in the samples as levels up to 10% of the total polypeptide content. Further details on the characterization of the impurity by MS are available in the Supplementary Information.

**Figure 1.**
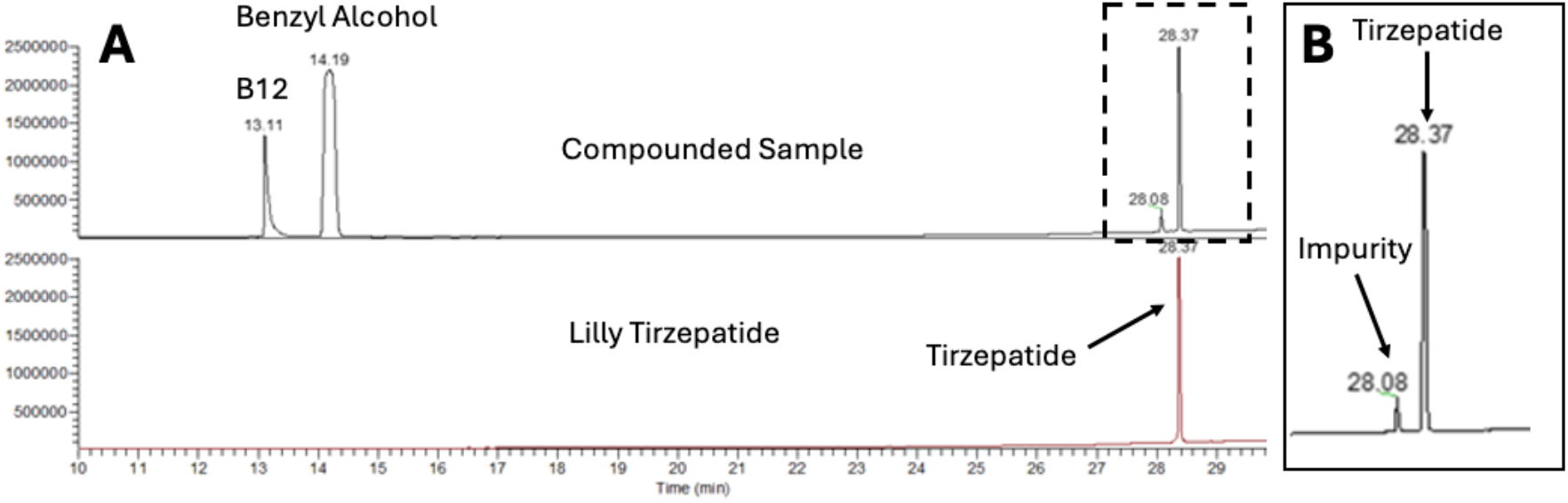

NMR experiments were undertaken to gain further insight into the interaction between tirzepatide and B12 in the solution phase at physiological pH and to investigate whether the interaction resulted in structural changes to the tirzepatide molecule. Both 1D and 2D NMR experiments demonstrated significant changes in the spectra of both tirzepatide and B12, indicating that tirzepatide and B12 interacted in the solution phase.

Figure 2 demonstrates significant changes in the NMR lineshape, intensity, and chemical shifts of the B12 signals in the presence of tirzepatide. Modest changes in the lineshape of tirzepatide signals were also observed. As tirzepatide exists as a trimer in solution, this is an expected result due to the differences in the molecular weight of a tirzepatide trimer (MW ∼ 13,000 Daltons) and B12 (MW ∼ 1,300 Daltons), as one would expect to see more significant changes in the B12 spectrum. These results are consistent with binding of B12 to tirzepatide in solution.

**Figure 2.**
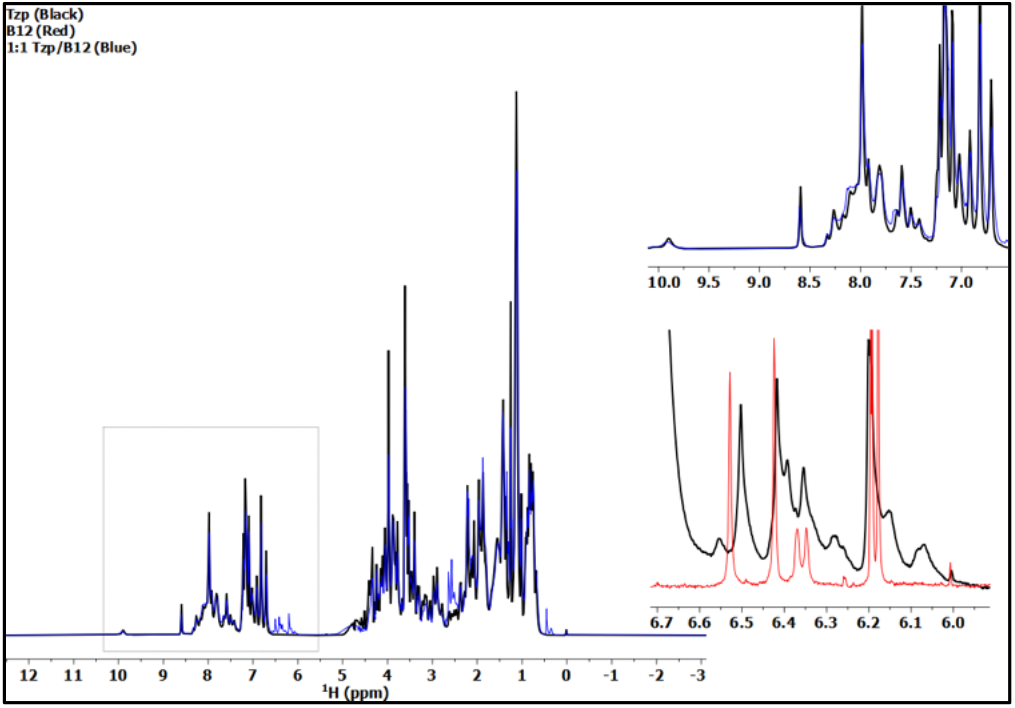
1D diffusion filtered proton NMR data indicating the presence of an interaction between tirzepatide and B12. Black traces are from tirzepatide alone, red from B12 alone, and blue from 1:1 mixture of tirzepatide/B12.

The two-dimensional proton-carbon heteronuclear single-quantum coherence NMR experiments (^1^H-^13^C HSQC) (Figure 3) indicated the presence of new peaks in the 1:1 tirzepatide/B12 sample that likely arise from alternate states of B12 in solution due to its interaction with tirzepatide. In addition, significant changes in lineshape and peak positions of the B12 resonance were observed, consistent with an interaction between B12 and tirzepatide Figure 4 shows results from the 2D ^1^H-^15^N HSQC NMR to study the impact on resonances in the polypeptide amide backbone region of tirzepatide. These experiments are generally considered the “fingerprint” for proteins and peptides, as the chemical shifts and pattern of resonances in this region of the NMR spectrum are generally indicated of the secondary and tertiary structure of the molecule. In drug discovery, the 2D ^1^H-^15^N HSQC experiments are the default NMR study used to probe the impact on the structure of a peptide or protein upon binding of a drug molecule. Changes in this region of the spectra are generally considered to represent changes in the secondary and/or tertiary structure of the molecule. The spectra in Figure 4 clearly show perturbations in the 2D spectra of both molecules, indicating changes in the structural properties of tirzepatide and B12 upon binding of B12 to tirzepatide.

**Figure 3.**
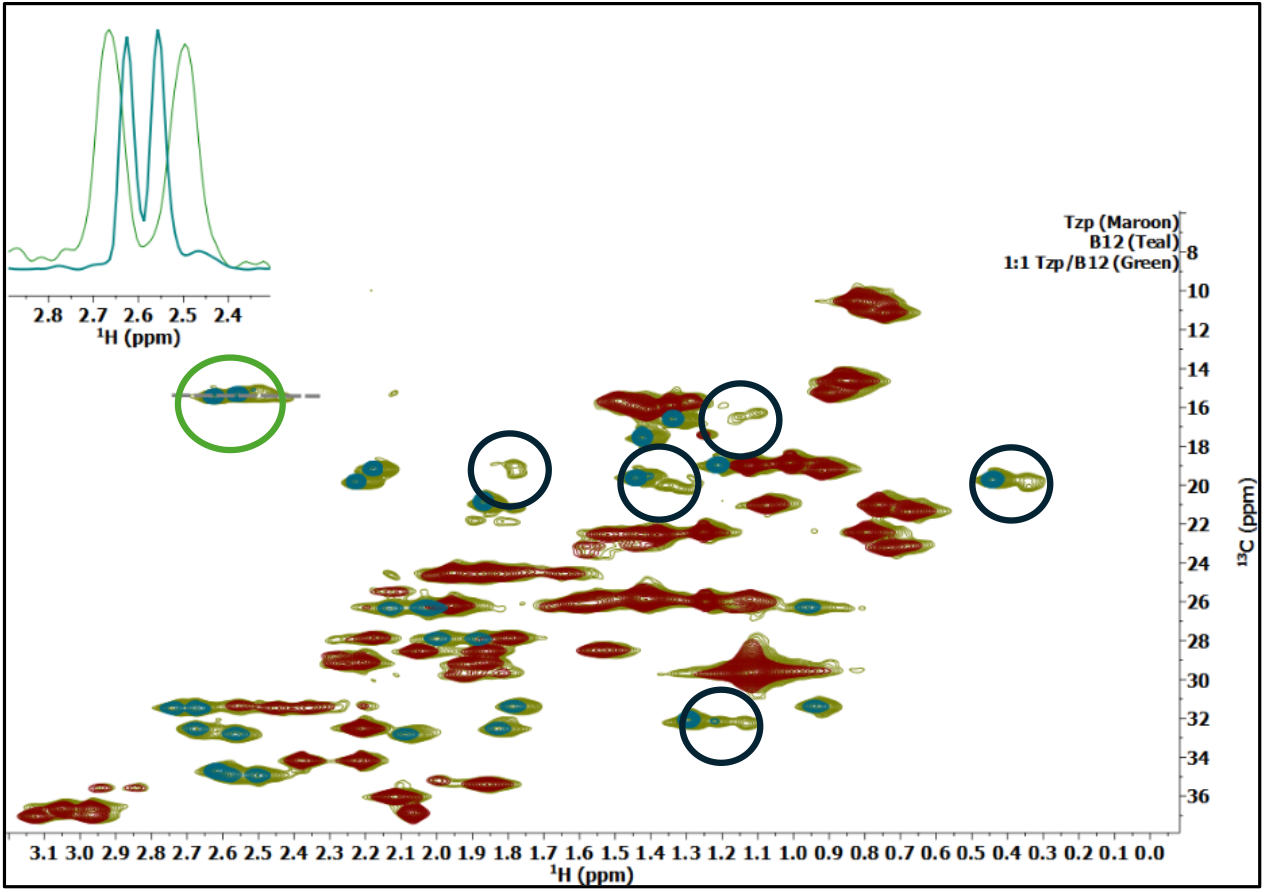
2D ^1^H-^13^C HSQC experiments indicating an interaction between B12 and tirzepatide. Spectra in maroon are from tirzepatide alone, teal are from B12 alone, and green are from 1:1 mixture of tirzepatide/B12. Notice that new green peaks show up near teal resonances, consistent with alternate states of B12 in the presence of tirzepatide. The inset on the upper left is a 1D trace of the peak circled in green and demonstrates an example of the significant changes in lineshape and chemical shift of the B12 resonances.

**Figure 4.**
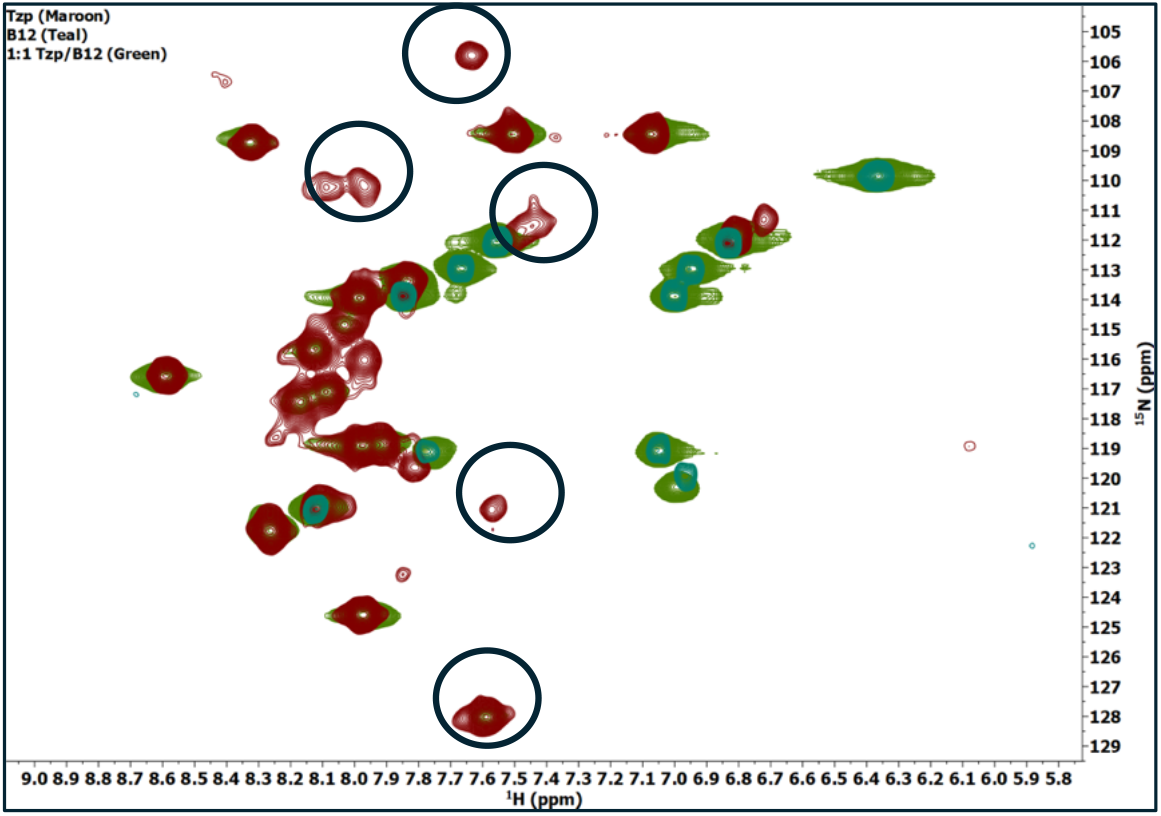
2D ^1^H-^15^N HSQC NMR experiments indicate changes to the structure of tirzepatide upon binding of B12. Maroon peaks are from tirzepatide alone, teal are from B12 alone, and green are from 1:1 mixture of tirzepatide/B12. The disappearance of some of the maroon and green peaks is indicative of binding to B12 and changes in structural properties of tirzepatide.

Lastly, the samples were analyzed by NMR experiments that examine diffusion rates of the molecules in solution. These types of experiments are used to probe conformational changes of molecules under various conditions. Generally, slower diffusion rates correspond to a larger molecular weight or hydrodynamic size. Additionally, these experiments report on the NMR relaxation rates of the molecules in solution. Increases in apparent molecular weight generally result in increases in T1 relaxation rates and decreases in T2 relaxation rates. The T1/T2 ratio is a commonly used metric to determine conformational and/or molecular size changes in solution. Increases in the T1/T2 ratio indicate an increase in apparent molecular size and conformational changes in the molecule.

Table 1 highlights the results from the diffusion NMR studies and demonstrates a decrease in the diffusion rate for both B12 and tirzepatide in the 1:1 samples. Further, corresponding increases in T1 relaxation and decreases in T2 relaxation lead to significant increases in the T1/T2 ratio of B12 in the presence of 1:1 tirzepatide and smaller increases in the T1/T2 ratio of tirzepatide in the presence of 1:1 B12. These measures suggest changes in the molecular conformation of each molecule. The changes in B12 are more prominent because B12 (MW∼1,300 Dalton) experiences a much larger increase in apparent molecular weight/size upon binding to tirzepatide (MW ∼13,000 Dalton in trimeric form). Similarly, tirzepatide experiences a decrease in its diffusion rate (e.g., tumbling slower in solution) and also an increase in the T1/T2 ratio. This increase is expected to be smaller, as the apparent increase in molecular size upon binding of B12 is smaller relative to what is experienced by the B12 molecule.

**Table 1.** Diffusion rates and relaxation times of B12 with and without tirzepatide and of tirzepatide with and without B12. Decreased diffusion rates are inversely correlated to molecular size and increases in T1/T2 ratio correlated with increased apparent MW of the tirzepatide/B12 complex.

|  | Diffusion<br>E<sup>-10</sup> m<sup>2</sup>/s | T1 (s) | T2 (s) | T1/T2 |
| --- | --- | --- | --- | --- |
| B12 only | 3.87 | 0.50 | 0.157 | 3.15 |
| B12 with 1:1<br>tirzepatide | 3.37 | 0.95 | 0.077 | 12.32 |
| tirzepatide only | 1.25 | 0.87 | 0.026 | 33.86 |
| tirzepatide with 1:1<br>B12 | 1.01 | 0.96 | 0.023 | 42.11 |

Taken together, these data demonstrate that hydroxocobalamin (vitamin B12) binds to tirzepatide under conditions similar to the tirzepatide drug product formulation and that this interaction produces structural changes in both molecules. The resulting tirzepatide–B12 adduct was present at substantial levels in compounded tirzepatide products containing hydroxocobalamin and methylcobalamin, both analogs of B12. The functional impact of this interaction on tirzepatide’s pharmacology, including binding to GIP and GLP-1 receptors, as well as its safety and efficacy, remains unknown.

## Discussion

The NMR data confirmed the presence of structural perturbations in tirzepatide when complexed with B12, suggesting that receptor binding may be perturbed, potentially altering efficacy or otherwise changing the drug’s pharmacologic profile. The larger molecular size of the tirzepatide–B12 adduct, as well as its altered structure, may also affect absorption, distribution, metabolism, and excretion, leading to potentially unpredictable pharmacokinetics. Additionally, protein-based drugs that are modified with other moieties—especially those that increase molecular size—can exhibit increased immunogenicity, raising concern that the tirzepatide–B12 adduct could increase the risk of unwanted immune responses [9].

To our knowledge, no studies, in animals, in humans, or in the lab have been conducted to assess the safety or potential toxicity of the tirzepatide–B12 impurity. Without such data, the impact of the impurity on efficacy, toxicity, and long-term outcomes for patients taking these compounded combination drugs is unknown and may be substantial.

### Patient Safety Risk and Broader Quality Concerns Associated with Compounded Tirzepatide

In the fall of 2025, it was reported that 80% of all compounded weight-loss drugs were combinations of tirzepatide or semaglutide with small, sub-therapeutic amounts of B vitamins or other amino acids [10]. Our finding of a previously uncharacterized impurity present at substantial levels in compounded tirzepatide mixed with B12 (a tirzepatide–B12 adduct)—which is absent from FDA-approved tirzepatide—represents an uncharacterized molecular species with an unknown safety profile and uncertain implications for safety, efficacy and immunogenicity. Its detection in compounded combination products represents a serious public health concern with implications for patients, healthcare providers, and regulators.

The tirzepatide–B12 adduct impurity appears to be one manifestation of broader quality and safety problems observed in compounded tirzepatide and other compounded products [11]. Compounded tirzepatide has been associated with a number of concerning quality issues, including contamination with bacteria or endotoxin, subpotent products, superpotent products, unlabeled active ingredients, active ingredients from unregistered suppliers, nonstandard excipients, and storage conditions that may compromise stability. Indeed, over 70,000 vials of compounded GLP-1 and GIP/GLP-1 products have been recalled since 2023, mostly due to lack of sterility [12]. Pharmacovigilance data from FAERS and FDA safety communications document substantial numbers of reported adverse events, including serious events requiring hospitalization and death, with use of compounded semaglutide and compounded tirzepatide products.

Further, misleading promotional practices for untested combinations of compounded tirzepatide with B12, other vitamins, and other substances are prevalent, particularly online [13]. These compounded combination products are marketed and dispensed to patients without adequate testing to understand the potential for interactions or to identify and qualify impurities, and without any sort of evaluation for safety or effectiveness.

The data presented provide a timely and clinically consequential warning about this increasingly common practice. Healthcare professionals should understand that these untested combinations offer no demonstrated benefit for patients versus FDA-approved tirzepatide yet present new potential risks through the formation of an adduct present at substantial levels (up to ∼10% of total protein content) as well as with broader concerns around product quality. Exposing patients to uncharacterized impurities without supporting clinical or toxicological data introduces unknown short- and long-term risks, underscoring the longstanding safety concerns associated with unapproved prescription drugs. Likewise, patients should understand that compounded tirzepatide may differ chemically and clinically from FDA-approved products and should therefore be informed that this impurity has been identified, and that quality problems with compounded drugs may reduce clinical benefit or increase the likelihood of adverse events.

### Regulatory Enforcement

Changes in enforcement and the regulatory framework are imperative to ensure that patients receive appropriate medications that have been demonstrated to be safe and effective through clinical trials and that they do not receive mass-compounded drugs under the guise of “personalization,” which circumvents federal restrictions on compounding without providing any sort of true clinical benefit (e.g., patients with B12 deficiency can be treated for that condition without creating an untested combination drug) and exposing them to new, unnecessary risk. Furthermore, the FDA approval framework exists to promote innovation while ensuring that products reaching patients are safe and effective. Allowing mass compounding outside this framework undermines both patient safety and the incentives that support innovative and generic drug development.

Several actionable improvements could materially reduce patient risk while bringing greater clarity to when compounding is appropriate—and when it becomes *de facto* drug manufacturing outside the FDA drug-approval framework. A core safety priority is to fully operationalize the compounding provisions of the Drug Quality and Security Act of 2013 (DQSA), many of which remain unimplemented more than twelve years later—including the demonstrable difficulties in compounding lists, the statutory limit on the volume of anticipatory compounding, and the statutory limit on interstate shipments of compounded drugs. Delay in implementing these and other DQSA provisions have created conditions that enable mass manufacturing under the guise of compounding and arguably pose an even greater threat to patient safety than those that existed at the time of the New England Compounding Center disaster [14,15].

Patient safety would also be strengthened by clarifying regulatory expectations for compounded copies of commercially available, FDA-approved medicines. Regulators need to clarify that minor or pretextual changes do not and cannot justify the mass-production of unapproved drugs. Small, clinically inconsequential changes to compounded products, like fixed-dose mixtures such as tirzepatide plus B12 and other analogs, introduce foreseeable and avoidable drug-interaction risks. This practices must be halted in order to protect patients unapproved products that are marketed outside the safeguards of the FDA-approval framework.

Pharmacovigilance and supply-chain transparency also remain critical gaps. Presently, mass-compounded drugs prepared in state-licensed pharmacies are not subject to adverse event reporting or any other federal pharmacovigilance requirement. At the same time, patients, clinicians, and often the pharmacies themselves lack visibility into API sourcing and emerging safety signals. Implementable changes include clearer documentation of API supplier qualification on certificates of analysis and a requirement for timely adverse-event reporting by entities that dispense compounded injectables—including telehealth-affiliated channels.

Finally, patient safety is undermined by misleading marketing claims that describe compounded combinations as “personalized,” “stronger,” or “safer” despite being mass-produced products that have not gone through clinical testing or regulatory review. Enforcing existing laws against misleading advertising and promotion for compounded drugs would reduce misinformation-driven demand and help prevent patients from unnecessarily receiving unapproved drugs.

## Conclusions

This previously uncharacterized impurity present at substantial levels in compounded tirzepatide mixed with B12 (a tirzepatide–B12 adduct)—which is absent from FDA-approved tirzepatide—represents an uncharacterized molecular species with an unknown safety profile and uncertain implications for safety, efficacy and immunogenicity. Its detection in compounded combination products represents a serious public health concern with implications for patients, healthcare providers, and regulators. Significant regulatory action is warranted to protect patient safety and changes in regulatory enforcement and full implementation of the DQSA is needed to restrict the mass manufacturing and interstate distribution of untested and unapproved drugs.

## Data Availability

All data produced in the present study are available upon reasonable request to the authors

