## Supplementary Information for "A Novel, Widespread Impurity in Mass-Compounded Tirzepatide/B12 Products: Patient Safety Implications"

**Supplemental Content**

**Experimental**

**Liquid Chromatography-Mass Spectrometry Analysis**

Samples were dissolved or diluted with diluent (5mM sodium phosphate, 140 mM sodium chloride, pH 7) to obtain a compounded tirzepatide concentration of approximately 1 mg/mL prior to analysis. Ultra performance liquid chromatography-mass spectrometric (UPLC-MS) analyses for sample characterization were carried out on a Waters ACQUITY UPLC H-Class system consisting of a Binary Solvent Manager, Sample Manager, PDA Detector, and 30-cm Column Heater with active solvent pre-heater using a Phenomenex Aeris peptide XB-C18 column. Mobile phases consisted of 0.1% trifluoroacetic acid (TFA) in water and 0.1% TFA in acetonitrile (ACN). The method utilized a 40 min elution time at a flow rate of 1.2 ml/min.

Mass spectrometry (MS) data were collected using a Thermo Orbitrap Exploris 480 with a heated-electrospray ionization (H-ESI) source, operating in positive-ion mode. Full-scan MS experiments were carried out at an orbitrap resolution of 120000, with other scan parameters optimized to target compounds of interest. Data-dependent MS/MS (ddMS2) experiments used an isolation window of 2 (m/z), an orbitrap resolution of 45000, and stepped higher-energy collisional dissociation (HCD) energies of 20%, 30%, and 40% for all analyses. Data were compared to an internal tirzepatide reference standard.

UPLC analysis of the sample revealed that the primary peak had a retention time consistent with FDA-approved tirzepatide per method criteria (see Figure 1 in manuscript).

Subsequent analyses were conducted to verify the identity of the unknown species. Comparison of the MS fragmentation pattern between tirzepatide and the peak 3 impurity showed the absence of the new species in native tirzepatide (Figure S1).

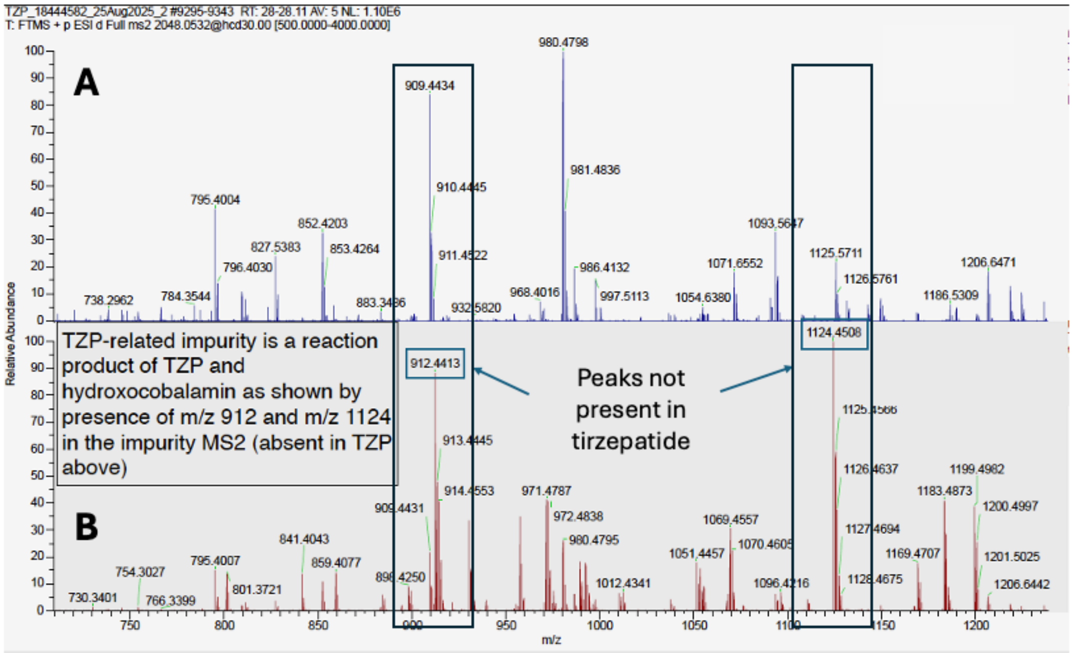


**Figure S1**. Comparison of ions present in native A) native tirzepatide and B) compounded tirzepatide + B12. Notice that the peaks at 912 and 1124 in panel B are absent in panel A, indicating the presence of a new species.

Accurate mass spectrometry analyses demonstrated that the new species possessed an intact mass of 6,138.07 Daltons (Figure S2).

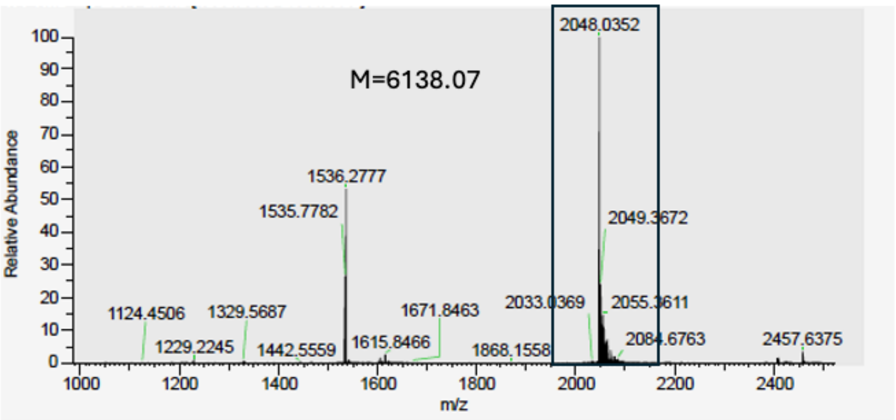


**Figure S2**. Full Scan Mass Spectrometry analysis of impurity peak. The ion at 2048.0352 is the +3 charge state of the tirzepatide-B12 adduct

The exact mass (theoretical monoisotopic mass) of hydroxocobalamin ($C_{62}H_{89}CoN_{13}O_{15}P$) is 1,345.57 Daltons, while the exact mass of tirzepatide ($C_{225}H_{348}N_{48}O_{68}$) corresponds to 4,810.52 Daltons. The mass of the impurity peak corresponds to 6,138.07 Daltons, which differs from intact tirzepatide by 1,327.55 Daltons. This indicates that the unknown species is a tirzepatide-cobalamin adduct that has formed through a likely ligand replacement reaction that resulted in the loss of a water molecule with a mass of 18 Daltons. Figure S3 is a schematic of the reaction that produces the tirzepatide–B12 adduct with mass balances noted.

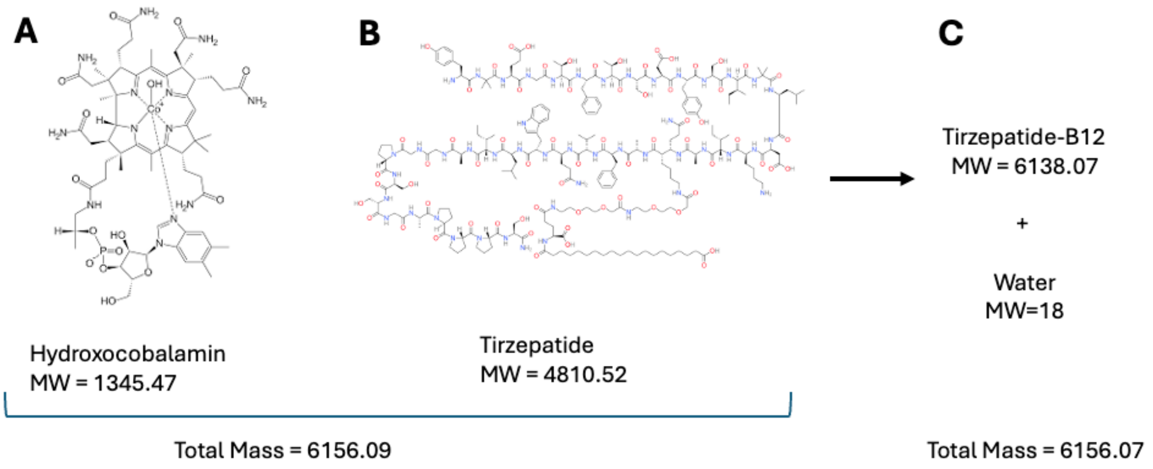


**Figure S3**. (A) Structure of hydroxocobalamin (MW 1345.47), (B) Structure of tirzepatide (MW 4810.52) and (C) reaction product of tirzepatide-B12 complex (MW = 6138.07) plus water (MW = 18).

**Nuclear Magnetic Resonance Experiments**

1D diffusion-filtered data were acquired using a pulse-field gradient stimulated echo experiment with bipolar gradients using 4 steady state scans, 512 acquisition scans, a 1 s recycle delay, a 0.2 s diffusion delay, a 0.7 ms gradient pulse applied at a strength of 45.7 G/cm, a spectral width of 20 ppm with 16384 points/FID, and a transmitter offset of 4.7 ppm. 2D $^{1}H$-$^{13}C$ ($^{1}H$-$^{15}N$) correlation spectra were acquired using the XLALSOFAST-HMQC experiment using 64 (steady state scans, 128 (256) acquisition scans, a 0.5 s recycle delay, forward and reverse INEPT transfer delays of 1.7 (2.4) ms, and spectral widths of 40 (30) ppm and 16 (16) ppm with 256 (128) points and 2048 points centered at 20 (117) ppm and 4.7 (4.7) ppm in the F1 and F2 dimensions respectively. Pseudo 2D diffusion data were acquired using a pulse-field gradient stimulated echo experiment with bipolar gradients using 32 steady state scans 64 acquisition scans, a 1 s recycle delay, a 0.2 s diffusion delay, a 1 ms gradient pulse, a spectral width of 20 ppm with 16384 point/FID and a transmitter offset of 4.7 ppm. Gradient strengths used were 17.6, 24.1 29.2, 33.6, 37.4, 40.9, 44.1 and 46.1 G/cm. Pseudo 2D T1 and T2 relaxation data were acquired using a custom sequence based on the 1D diffusion-filtered experiment (Bradley, S. A.; Jackson, W. C.; Mahoney, P. P. Measuring Protein Concentration by Diffusion-Filtered Quantitative Nuclear Magnetic Resonance Spectroscopy. Analytical Chemistry 2019, 91 (3), 1962-1967. DOI: 10.1021/acs.analchem.8b04283.) using 16 steady state scans 64 acquisition scans, a 1s recycle delay, a spectral width of 20 ppm with 16384 points/FID and transmitter offset of 4.7 ppm. T1 relaxation delays used were 0.1, 0.2 0.4 0.8 1.2 and 1.6 s. T2 relaxation delays used were 4, 8, 16, 32, 48 and 64 ms.
